# Passive sensing with psyche: utilizing data from wearable technology to predict emotion states

**DOI:** 10.1101/2025.02.13.25322254

**Authors:** Helmet T. Karim, Ajmail Matin, Mayank Goel

## Abstract

Depression and anxiety are some of the most common mental health disorders in the world contributing to significant morbidity and mortality. Past treatments have focused primarily on treating depression and anxiety. However, there is an urgent need to detect chronic stress states and potentially intervene using just-in-time personalized interventions. Modern technology has revolutionized our ability to passively measure various biological and physiological signals. In our daily lives, we generate significant amounts of electronic data from our phones, wearable technology, watches, and even computers and cars. In this analysis, we focus on using wearable data from FitBit to passively predict daily mood states (e.g., sad/tense/anxious vs. happy). We use daily FitBit data from 38 participants and ~1200 days of data to predict mood states (e.g., sad/tense/anxious vs. happy) on a day-to-day basis using an elastic net regression machine learning algorithm. We were able to accurately predict these states using a cross-validated machine learning algorithm and identified features predictive of each of the mood states. In this proof-of-concept analysis, we show that predicting daily mood states is feasible and may help to not only detect daily mood states but also improve passive awareness and deliver just-in-time interventions.

## Introduction

Depression and anxiety are the most prevalent mental health disorders in the world^1^. These disorders are associated with significant morbidity, mortality, and reduced quality of life^2^. While many studies have focused on the treatment of these disorders, far fewer have focused on the detection of chronic stress states and potential intervention prior to a mental health diagnosis. For example, mindfulness meditation can reduce depressive symptoms even in individuals with low or moderate symptoms^3-5^. Mental hygiene are practices that promote mental health and improve overall well-being that aim to prevent mental health disorders like depression and anxiety. For example, one study found that mindfulness practices can improve negative repetitive thoughts, including anxiety, worry, and rumination^5^. One major challenge is to improve self-awareness of these stressful states or help individuals to become better aware of these states. One potential avenue is ‘passive sensing.’

Passive sensing is the ability to detect mental health states using wearable or passively collected data along with machine learning. Technology like smartwatches, phones, and other biosensors have revolutionized the type of data that we can collect on individuals^6^. They are widely popular and ubiquitous, with an impressive ability to capture temporal and daily patterns of various biological and physiological signals. There are several reviews and studies that have shown that this type of prediction is feasible and accurate^7,8^. Various features like sleep, physical activity, and heart rate have been used to predict mood disorders^9-11^. These approaches may be able to improve our ability to detect and then potentially intervene actively.

One major limitation is that many of these past studies have focused on predicting measures like PHQ9, which measures average depressive symptoms over a period of time. One of the major drawbacks to this is recall bias and a hyperfocus on severe depressive symptoms. In this analysis, we focus on predicting day-to-day mood states, which may allow us to detect daily stress and potentially deliver just-in-time interventions to counteract these negative mood states. We analyze data from the LifeSnaps study that measured daily ecological momentary assessments (EMA) along with daily FitBit data. We use this data to predict sad/anxious/tense compared to happy states and identify features predictive of these states.

## Methods

### Study Design and Participants

We used data from the LifeSnaps study sourced from Kaggle^12^. The details of this study are provided elsewhere, but we provide a brief overview here^12^. Participants were recruited from four sites and was approved by the Institutional Review Board (IRB) of the Aristotle University of Thessaloniki (Protocol Number 43661/2021). There was a total of 71 participants (convenience sample) recruited. Participants needed to a mobile phone and were at least 18 years old and were willing to wear a sensor to collect Fitbit data (https://www.fitbit.com/global/us/products/smartwatches/sense) for the duration of the study. In addition to Fitbit data, participants completed daily EMA as well as other surveys. For the purposes of the current analysis, we only analyzed daily measures.

### Fitbit data

This data was collected with a FitBit Sense device. This was collected in a broad setting without any restrictions, e.g., collected passively. This device has a 3-axis accelerometer, gyroscope, altimeter, built-in GPS, optical heart rate tracker, skin-temperature device, ambient light detector, and multipurpose electrical sensor^12^. For our purposes, we used a number of features that were converted to daily measures including nightly temperature, non-rapid eye movement hours, root mean square of successive differences (RMSSD) of heart rate, distance traveled that day, average beats per minute, lightly active minutes, moderately active minutes, very active minutes, sedentary minutes, average resting heart rate, minutes asleep, minutes awake, sleep efficiency, and number of steps.

### Ecological Momentary Assessments (EMA)

Participants also completed daily EMA’s to assess various features related to goals, mood, and location. In this analysis, we primarily focused on mood, in particular we focused on items tense/anxious, happy, and sad. There were other mood ratings, however we decided to focus on affective ratings rather than the others (neutral, alert, rested/relaxed, and tired).

### Final Analytical Sample

As we were primarily interested in passive sensing, where we predict their mood state based on wearable data, we were only interested in analyzing data available on days where these assessments were completed and available. Initially, this dataset including 7,410 days of observation across participants. When we dropped rows with unavailable data (across any of the features), we resolve to a final analytical sample of ~1,200 days. This ultimately included data on 38 participants with totally complete data. We used a multinomial elastic net regression to predict days where participants reported being happy (276 days), tense/anxious or sad (313 days), or neither (578 days).

### Elastic Net Regression

We used the eNetXplorer^13^ to predict the multinomial label using a 5-fold cross-validation optimizing over average accuracy. Elastic net regression combines the L1 norm penalty (|ß|) of LASSO (least absolute shrinkage and selection operator)^14^ with the L2 norm penalty (ß^2^) of ridge regression^15^ by including a parameter α such that α = 1 reduces to LASSO, while α = 0 reduces to ridge regression. It still has the penalty for each subsequent non-zero ß value called λ. For each model, we optimize over various values of α=0-1 every 0.05 and 100 values of λ. We repeated this 500 times. We permuted the outcome and repeated this process with 250 permutations to compute null bounds on the prediction accuracy. This approach can estimate a *p*-value of the models as well as the *p*-value for each individual independent variable^13,16,17^.

## Results

We found that our model performed accurately with approximately 70% accuracy, p<0.001 (α=0, λ=53.9). We were able to accurately predict each label and show features that predict each label indicating the feature coefficient and p-value (table 1).

**Table 1.** Predictors of each event – features are shown on the left while each individual event (or lack of) and their predictors along with average feature coefficient and p-value is shown. Features are ordered by average rank order based on p-values. Values in bold indicate significant predictors of each component.

|  | No Event |  | Happy |  | Sad or Tense/Anxious |  |
| --- | --- | --- | --- | --- | --- | --- |
| Measure | coefficient | p-value | coefficient | p-value | coefficient | p-value |
| Steps | -0.00041 | 0.1224 | **0.00084** | **0.0002** | -0.00043 | 0.0689 |
| Distance Traveled | -0.00041 | 0.1241 | **0.00084** | **0.0003** | -0.00043 | 0.0722 |
| Nightly Temperature | -0.00026 | 0.3371 | **-0.00073** | **0.0012** | **0.00099** | **0.0000** |
| Very Active Minutes | 0.00028 | 0.2980 | **0.00055** | **0.0167** | **-0.00082** | **0.0005** |
| Minutes Awake | **0.00076** | **0.0041** | -0.00038 | 0.0999 | -0.00038 | 0.1082 |
| Moderately Active Minutes | -0.00040 | 0.1395 | **0.00076** | **0.0006** | -0.00037 | 0.1250 |
| Sleep Efficiency | **-0.00062** | **0.0212** | -0.00011 | 0.6211 | **0.00073** | **0.0023** |
| Lightly Active Minutes | **-0.00065** | **0.0145** | 0.00008 | 0.7384 | **0.00058** | **0.0162** |
| NREM Hours | **0.00053** | **0.0470** | **-0.00052** | **0.0231** | -0.00001 | 0.9609 |
| RMSSD | -0.00020 | 0.4693 | **0.00048** | **0.0365** | -0.00028 | 0.2355 |
| Sedentary Minutes | 0.00015 | 0.5795 | **-0.00047** | **0.0392** | 0.00032 | 0.1812 |
| Resting HR | 0.00037 | 0.1621 | **-0.00047** | **0.0426** | 0.00010 | 0.6860 |
| Avg BPM | 0.00031 | 0.2476 | -0.00014 | 0.5553 | -0.00018 | 0.4637 |
| Minutes Asleep | 0.00002 | 0.9454 | 0.00022 | 0.3362 | -0.00024 | 0.3199 |

The following features predicted the happy label (ordered by p-value): number of steps, distance traveled, moderately active minutes, nightly temperature, very active minutes, NREM hours, RMSSD, sedentary minutes, and resting HR.

The following features predicted the sad/tense/anxious label (ordered by p-value): nightly temperature, very active minutes, sleep efficiency, and lightly active minutes.

The following features predicted the non-happy or sad/tense/anxious label (ordered by p-value): minutes awake, lightly active minutes, sleep efficiency, and NREM hours.

## Discussion

In this proof-of-concept analysis, we found that we were able to passively detect emotion states using just data from wearable technology. We were able to do so with relative accuracy and identify predictive features for each of the various states. This type of passive detection can improve emotional awareness by identifying and helping individuals gain passive awareness of stressful states (e.g., tense/anxious or sad mood states). This can not only potentially enhance self-awareness and improve emotional intelligence but can also identify important states for intervention. Through passive detection we can identify stressful states and potentially deliver just-in-time interventions to help improve mood and prevent or reduce the likelihood of subsequent mental illness.

Our model was able to predict accurately using data from just the FitBit sense system. We call this passive sensing as we are able to detect mood states using passively measured features which are now widely available. In addition, we showed the capacity to passively detect happy and sad/anxious states on a day-to-day basis using a more richly phenotype dataset. Despite this, we were only able to achieve 70% accuracy in part potentially due to the more limited nature of the data as well as the more limited nature of the daily assessments as only ~15% of the days with complete FitBit data also had EMA completed.

Past studies have primarily focused on predicting depression and anxiety on average using assessments like PHQ-9 or GAD-7, respectively, but these measures estimate average levels over typically one or more weeks in a retrospective fashion^18,19^. One such study used data from 10k participants’ FitBit data to predict these measures with relative accuracy^19^. Our model focuses on predicting daily mood states, which may be able to be embedded along with digital interventions to detect stress, deliver interventions accurately, and improve overall well-being.

There are some major limitations primarily that this is a proof-of-concept approach to show that daily sensing of mood states is feasible. Despite our relatively modest accuracy, we were able to achieve this with limited data available. The overall sample was relatively healthy without severe mental health disorders – this may be critical for translating to not just the real-world environments but also into clinical environments.

We were able to accurately predict daily sad/anxious/tense compared to happy states using wearable sensor data. This type of passive sensing allows for collection of data from everyday wearables that have become ubiquitous to everyday life and detecting stressful states without any user input. In the future, we hope to expand this technology to allow for individualized models that use a person’s own data to predict their stressful mood states using a personal model. This can not only potentially better predict these states but also has the capacity for identifying individualized features that can be optimized for intervention.

### Psyche: Transforming data into wellness

Using artificial intelligence (AI) may help transform data collected passively into wellness. We can apply passive sensing technology to real-world mental health care by integrating models into various systems including electronic health records (EHR) and telehealth platforms. Through passively collected data, we can provide real-time insights into patient mood patterns, offering objective measures to complement subjective self-report and period evaluations which can reduce patient burden. We can build AI powered systems that can detect periods of distress early, improve patient self-awareness, and provide actionable insights that minimize workflow disruptions and optimizing and personalizing treatment for patients. In addition to detection and awareness, we can provide digital just-in-time interventions, such as mindfulness exercises or breathing prompts, triggered by real-time physiological and behavioral signals. Future studies can help assess impact on these types of systems on various outcomes including symptoms and patient/physician engagement. This approach may have potential to enhance physician and therapist decision-making and improve patient outcomes, offering a scalable model for proactive, data-driven mental health wellness.

## Data Availability

All data is publicly available on kaggle: https://www.kaggle.com/datasets/skywescar/lifesnaps-fitbit-dataset.

## Acknowledgements

We would like to acknowledge and thank the LifeSnaps data investigators for making this data publicly accessible.

